# Cannabis, identity, and attitudes: a qualitative study in adolescents who do and do not use cannabis

**DOI:** 10.1101/2022.08.15.22278796

**Authors:** Christiana Karashiali, Will Lawn, Kat Petrilli, Claire Mokrysz, Georgia Black

## Abstract

**Background and objective:** Cannabis use during adolescence is common. Cannabis use and identity are thought to influence one another. This study aimed to examine what London-based adolescents (aged 16-17 years) think about cannabis use, its relationships with identity, and its benefits and harms.

**Method:** Three semi-structured focus groups interviews were conducted, two with adolescents who use cannabis (n=3 and n=5) and one with adolescents who do not use cannabis (n=6). Participants also completed a drug-use questionnaire.

**Results:** Thematic analysis (TA) revealed four identities. Two identities emerged from both groups: ‘The person who uses cannabis is chilled’ and ‘The person who uses cannabis is sometimes ostracised’. Two identities emerged from the group of adolescents who used cannabis: ‘The person who uses cannabis is an expert in risky things’ and ‘The person who uses cannabis is not addicted’. Skunk was identified as potentially more harmful than hash, but more powerful and pleasurable.

**Conclusion:** The findings provide insight into how cannabis use shape personal and social identity amongst teenagers in London in the late 2010s. Those who use cannabis described the benefits of cannabis, including socialising and for relaxation, and emphasised they are not addicted. Stigmatising and devaluing attitudes were held by some non-users about adolescents who use cannabis. Stereotypes seem to still exist, despite cannabis normalisation. Implications for research and policy are outlined.

## Introduction

Cannabis is the most commonly used internationally regulated drug, with 188 million global users in 2017 (UNODC, 2019). It is a regular feature of many adolescents’ lives, as approximately 20% of 15-year-olds in England had used cannabis in the past year (NHS Digital, 2018). Teenagers are more likely to use cannabis than adults (EMCDDA, 2015), therefore understanding adolescents’ thoughts and attitudes about cannabis use is important.

Cannabis use was historically characterised as a deviant behaviour (Becker, 1953) but has become ‘normalised’ in the last 30 years (Hammersley et al., 2001), as indicated by high use rates (Hathaway et al., 2011). Cannabis produces desirable short-term effects, such as relaxation and euphoria (Green et al., 2003). However, heavy, long-term use is associated with a small increased risk of mental health problems and mild neurocognitive impairment (Curran et al., 2016).

Adolescence is a crucial period for self and identity development, including physical, psychological and social changes (Marcia, 1987). These changes theoretically lead adolescents to ask ‘Who am I?’ (Erikson, 1968) and develop an integrated understanding of their identity. Whilst the process of identity development begins in childhood, it becomes a primary concern during adolescence and the identities created in childhood may be altered (Erikson, 1968).

Identity is conceptualized in two ways: self-identity, the primary and enduring views of one’s self-perception (Sparks, 2000) and group identity, a person’s sense of belonging to a specific group (Chen & Li, 2009). These are both thought to evolve during adolescence. Self- and group identity are related, as social identity theory (SIT) suggests that self-development occurs through interaction with similar and dissimilar groups (Tajfel, 1982).

Cannabis use can affect identity formation and vice versa (Lamb, 2011). Hammersley et al. (2001) suggest that cannabis use is an important aspect of many people’s individual and group identities. For example, Miller et al. (2016) reported that identifying with family and school groups, versus friend groups, is associated with lower odds of cannabis use. Weiss et al. (2011) found that adolescents with negative self-image were 1.5 times more likely to be users than those with a positive self-image. Also, qualitative studies have found that adolescents who use cannabis view it as less harmful than tobacco and alcohol (Akre et al., 2010; Menghrajani et al., 2005). A longitudinal qualitative study found a gradual evolution of attitudes towards cannabis from ages 12-13 to 16-17 years (Bilgrei et al., 2021). The study found that initially, cannabis was a symbol of addiction and marginalization, with anti-drug education sentiments regularly echoed and exaggerated. In later adolescence, teenagers expressed more nuanced opinions about cannabis and some viewed the associated risks as acceptable. A wider array of symbolic images were drawn from digital media and popular culture.

Despite the widespread use of cannabis among adolescents, qualitative research in this field is limited, especially in the UK. Qualitative research can give an insight into the complex nature of values, identities and sociocultural context of psychological phenomena (Banyard & Miller, 1998). Therefore, this study gave adolescents living in London a voice to describe how cannabis contributes to their lives and identities. As smoking high-potency cannabis (sinsemilla or ‘skunk’) is associated with a higher risks of mental health problems (Di Forti et al., 2019; Freeman & Winstock, 2015), we aimed to explore what adolescents think about different types of cannabis. Our research questions were:

- How do cannabis use and identity affect each other?
- What do adolescents who use cannabis think the benefits and harms of cannabis use are?
- What do non-using adolescents think about their cannabis using peers?
- What do adolescents think about different types of cannabis?

## Method

### Design

A qualitative study using focus group data to explore adolescents’ perspectives on cannabis use. The study was approved by the University College London (UCL) Ethics Committee with project ID 5929/003.

### Recruitment

Participants were identified from an existing database of participants and paid-for online adverts. We conducted a telephone screening to assess eligibility. Inclusion criteria were: (1) 16-17 years old, (2) fluent in English, (3) capacity to give informed consent, (4) (if a user) use cannabis one or more days per week, and (5) (if a non-user) never have used cannabis. Exclusion criteria were: (1) not use any other illicit drug daily or almost daily, (2) not receive treatment in relation to any mental health or substance use condition. Participants received £15 each, to cover their travel expenses.

### Participants

Fourteen participants took part, 5 males and 9 females. The focus groups were as follows: group 1 with teenagers who use cannabis (n=3; 1 male and 2 females); group 2 with teenagers who use cannabis (n=5; 3 males and 2 females); group 3 with teenagers who do not use cannabis (n=6; 1 male and 5 females).

### Procedure

#### Data collection

The study took place in at UCL. On arrival, informed consent was received from all participants.

### Measures

#### Interview schedule

An interview schedule, including potential questions and prompts was used. Each focus group began by participants drawing or writing anything that came to their mind on what a ‘stereotypical’ person who uses cannabis looks like. The researchers then initiated discussion by displaying the depictions and asking participants how they compared to themselves or their friends. Broad questions followed, such as ‘Tell me about a typical scenario when you/people you know tend to use cannabis’. Researchers encouraged discussion by using prompts, such as ‘Who’s there in this scenario?’. Towards the end of the focus group, participants were shown a video of young males talking about their experiences with cannabis, both positive and negative. The aim was to explore whether participants recognised any of the described consequences in themselves or peers, and to prompt discussion of other consequences they may have experienced. Each group lasted around 50 minutes. Conversations were recorded and professionally transcribed, in preparation for subsequent thematic analysis. All data remained confidential and participants were given a pseudonym.

#### Self-report questionnaires

Participants completed a questionnaire, detailing demographics and drug use history. The questionnaire assessed ever use and age of first use for alcohol and cigarettes, and for users also assessed age of first cannabis use. Additionally, users were asked their most common method of cannabis administration and cannabis type used, over the past three months. To indicate cannabis type, they chose between images depicting different cannabis types: skunk, hash and herbal.

Users also completed the Cannabis Self-Concept scale, a 5-item scale assessing explicit association between cannabis use and identity (Blevins et al, 2018). Participants indicated the extent to which they agree (1=Strongly disagree, 10=Strongly agree) with five statements (e.g., ‘Smoking cannabis is part of my self-image’).

Users also completed the Cannabis Use Disorder Identification Test-Revised (CUDIT-R), an eight-item self-report cannabis misuse-screening instrument (Adamson et al., 2010).

### Data analysis

Data were analysed using Thematic Analysis (TA) to investigate patterns across the data set (Braun & Clarke, 2006). TA was selected as it enables a systematic but loose structure for data analysis and an interpretation of participants’ thoughts and feelings in a realistic context. The analysis followed five phases: ‘data familiarisation’, ‘coding generation’, ‘looking for themes’, ‘reviewing and refining themes’ and ‘theme labelling’. Firstly, the transcripts were read multiple times to familiarise oneself with the data. Relevant extracts were then inductively coded, meaning that they were coded without aiming to fit them into a pre-existing coding structure, or the researcher’s preconceived ideas. Researchers aimed to invoke psychological terminology, whilst taking a critical realist epistemological position, taking the data at face value, but also bearing in mind that people formulate their own interpretation of the world. The codes were clustered into sub-themes, which were then grouped into themes. To ensure credibility, themes and subthemes were discussed and refined between two researchers (GB and CK).

## Results

### Sociodemographics

Participants ranged between 16.2-17.9 years of age, with a mean age of 17.2 years old. Sociodemographic characteristics are illustrated in Table 1.

**Table 1.** Summary of sociodemographic characteristics and drug use variables.

| Demographics | Participants (n=14) |  |
| --- | --- | --- |
|  | Non-users | Users |
|  | (n=6) | (n=8) |
| Age (years old) | 17.18 (0.68) | 17.18 (0.46) |
| Gender |  |  |
| Male | 1 | 4 |
| Female | 5 | 4 |
| Ethnicity |  |  |
| White English | 1 | 4 |
| White and Asian | 0 | 1 |
| White and Persian | 1 | 0 |
| African | 2 | 0 |
| American Australian | 0 | 1 |
| Black Caribbean/ Greek Cypriot | 0 | 1 |
| Pakistani | 1 | 0 |
| Turkish | 1 | 0 |
| Any other mixed | 0 | 1 |
| Cannabis frequency (days per week) over the<br>past 3 months |  | 4.64 (1.57) |
| Age when first tried cannabis |  | 14.57 (1.13) |
| Age when first began using cannabis every week |  | 16.04 (0.73) |
| Age when first tried alcohol | 15.68 (0.98) | 13.23 (1.04) |

Note. n= total number

### History of alcohol, cigarette and cannabis use

Over the past three months, five users mostly used skunk, two herbal and one hash. Users used cannabis at a frequency of 4.64 (SD=1.57) days per week. Participants had a mean CUDIT score of 16.38 (SD=4.69). Table 2 indicates the results from the Cannabis Self-Concept scale, with a mean total score of 4.05 (SD=0.72).

**Table 2.** Mean agreement scores (SD) for each item on the Cannabis Self-concept Scale (0 to 10).

| Statements | Mean agreement score |
| --- | --- |
| Total | 4.05 (0.72) |
| Smoking cannabis is part of my self-image | 3.88 (2.09) |
| Smoking cannabis is who I am | 3.13 (2.62) |
| Smoking cannabis is part of my personality | 3.63 (2.45) |
| Smoking cannabis is a large part of my daily life | 5.25 (2.17) |
| Others view smoking cannabis as part of my personality | 4.38 (1.80) |

### Findings from the TA

Four key identities were identified and discussed below.

#### Theme 1: The person who uses cannabis is chilled

Both groups described the ‘typical’ person who uses cannabis as being relaxed and ‘chilled’. Some celebrities were raised as examples of famous cannabis users. Many focused on external appearance and clothing:

> *Many associate cannabis with people who are urban…a lot of tracksuits and the example of Wiz Khalifa [Aleena, non-user]*.

Sophia, a non-user, highlighted that the choice of clothing could imply that the stereotypical cannabis user is inactive, in a denigrating way:

> *He’s wearing a puffer jacket and trackies, just sitting there, not doing much [Sophia, user]*.

Users also reflected that cannabis has a positive functional value in terms of relaxation and stress relief. By reducing stress, some also saw cannabis as a ‘social lubricant’:

> *I find it easier to talk to people and level with them [Laura, user]*.

Both groups portrayed the ‘typical’ cannabis user as being relaxed in both their demeanour and outward presentation. This was reported to have positive effects on stress and socialising. However, there were also judgmental insinuations of inactivity or laziness associated with this trait.

#### Theme 2: The person who uses cannabis is sometimes ostracised

Both groups articulated stereotypes of people who use cannabis being less serious and their behaviour sometimes even being immoral. This caused non-users to distance themselves from users, who experienced this as a form of social rejection.

For example, non-users mentioned that their using counterparts are less academically-focussed:

> *They’re normally the academically unserious…We’re in the library, they’re playing football while everyone’s working [Zehra, non-user]*.
>
> *They’re quite loud…very outspoken [Jordan, non-user]*.

In addition to unserious and immoral characteristics, non-users made comments about class. One non-user noted the perceived incongruence between being “upper class” and smoking cannabis:

> *There are some unserious people who smoke every weekend… I had some friends who were quite upper class, dressed nice, it didn’t fit their image, but they smoked [Alice, non-user]*.

In addition to classist stereotypes that relate to parental background, Essie (a non-user) held moralistic beliefs about users’ parents:

> *They have a ‘trap house’. A group of them all in one house. I’m thinking, ‘Where’s their parents?’ [Essie, non-user]*.

Some non-users tried to ensure that they did not become associated with users’ ‘devalued’ identity or become influenced by their behaviour, and were relieved that users tended to stay together:

> *I’m completely against it… It’s good because it keeps them together. [Aleena, non-user]*.

Whilst some non-users were happy to be friends with users, they explained that they might avoid larger groups of users:

> *One of my best friends smokes. I wouldn’t want to stop being friends with her because she smokes. That’s her decision and her life, but if she does a gathering with people who smoke, I won’t come [Amara, non-user]*.

Female users in particular were aware of being devalued and socially rejected. Some expressed their experience of rejection:

> *My friends didn’t think it was a good thing. They didn’t want to associate with that stuff, so they stopped talking to me. Then, I started talking to this girl I’m friends with now. She’s the only friend I have [Jessica, user]*.
>
> *A lot of them stopped talking to me. I now have a best friend who’s pretty much the same as me [Laura, user]*.

This suggests that as well as having cultural associations about academic achievement and class, adolescent women may be particularly vulnerable to judgment about cannabis use.

Users described choosing an encouraging environment within which they feel comfortable to smoke. Despite being rejected by their friends, they tried to find people who shared their attitudes to cannabis, instead of altering their behaviour to ‘fit the group’:

> *I smoke with close friends. I must be with people I can resonate with, who I’m on the same level with [Justin, user]*.

Social media platforms were perceived to have a significant role in how cannabis users related to each other, reinforcing attitudes and behaviours by publicly demonstrating their identity as a user:

> *I put weed in my story [Snapchat]. It’s a lot to do with pride. I don’t feel like cannabis is part of my identity but people probably view me as that guy, because I post it [David, user]*.

David expressed that it matters how others perceive him, but cannabis does not impact how he views himself. Rather than accepting a devalued identity, he related this to his passion for cannabis.

#### Theme 3: The person who uses cannabis is an expert in risky things

Adolescent users, despite being aware of cannabis risks, seemed to enjoy talking about their experiences of negative effects. Despite admitting that the effects were ‘bad’, there was humour in the re-telling:

> *When people are talking to me, I’m like, ‘mm, yes’ [laughter] but I’ve lost track of what they were saying five minutes ago*… *It’s really bad [Noah, user]*.

Several different negative effects were mentioned including Noah’s experience of memory loss or confusion, and some users experienced more severe effects, particularly from using strong cannabis:

> *I had 2 grams, the strongest cannabis…It was horrible! It felt like schizophrenia [David, user]*.
>
> *I had a panic attack. I couldn’t breathe. Another time I passed out about ten times in a row… I didn’t feel like a human; I felt like a worm [laughter] [Sophia, user]*.

Although Sophia was aware of the potential risks and relates this incident as a dramatic event, she also downplayed it by using humour and exaggeration. Users were aware of long-term harms associated with cannabis:

> *I’ve never felt like I’m getting close to schizophrenia or psychosis, but I know it’s there [Justin, user]*.

Users often referred to other users as examples of people whose mental health had deteriorated which caused them to be concerned for themselves. However, they had conflicting beliefs about their own ability to control the risks through their expertise in cannabis use and its associated dangers.

Examples of reference to their expertise included making informed decisions about the type of cannabis they use. For example, the use of hash could make them more relaxed, whereas they perceived skunk to be more risky:

> *From skunk, people can really go psychotic and get sectioned [Jessica, user]*.

Despite this knowledge about the risks of smoking skunk and its association with mental health problems, some users reported that they preferred skunk:

> *I like skunk more, it’s a special place in my heart [David, user]*.

Despite having unpleasant experiences and being aware of the risks, users enjoyed their expertise in the riskier aspects of cannabis use.

#### Theme 4: The person who uses cannabis is not addicted

Users emphasised their capacity to make active choices, as well as rejecting ‘addicted’ status. Users stated that it is their choice to smoke and something they enjoy doing:

> *You’re not really addicted, you feel like you just want to use it more [David, user]*.

David describes wanting to “use it more”, but his use of the word “you” instead of “I” could suggest that he presumes a shared view is held amongst other users. Similarly, Lee rejected the idea of being addicted:

> *It’s not even like an addiction. It’s just something you do*… *It’s not based on your personality or what you look like [Lee, user]*.

Users clearly perceived cannabis use to be a personal choice and a self-regulated behaviour. They saw themselves not as ‘addicts’ but rather ‘seekers of pleasure’. Participants talked about their ability to control how much they smoked, to reinforce that they were not addicted.

> *I’ve been smoking for two years. If I wanted to stop, I could [Laura, user]*.

Overall, users prioritised personal values to reflect how they see themselves. For them, smoking was a personal decision and they used it because they enjoyed it, whilst for non-users, cannabis abstinence was a moral decision.

## Discussion

This study captured a group of adolescent’s perspectives on cannabis use and explored it in the context of self and group identity formation. We recognised four key identities that were identified differentially by adolescents who do and do not use cannabis; ‘The person who uses cannabis is chilled’, ‘The person who uses cannabis is sometimes ostracised’, ‘The person who uses cannabis is an expert in risky things’ and ‘The person who uses cannabis is not addicted’. Adolescents who used cannabis mainly viewed their use of the drug as pleasurable, not-addicted, and serving important functions. Prejudiced attitudes were held about adolescents who use cannabis, by some of their non-using counterparts.

Our findings partially support the idea of the normalisation of cannabis use in mainstream youth culture in London, but also suggest a more complex picture in which using cannabis is still somewhat stigmatised (Hammersley et al., 2001). These results are reminiscent of a longitudinal qualitative study conducted with Norwegian adolescents (Bilgrei et al., 2021), suggesting that these attitudes are relevant in different countries and contexts. Many adolescents who do not use cannabis perceived those who do as ‘unserious’, as well as associating cannabis use with problematic class and gender-related stereotypes. Indeed, two participants proposed that cannabis should be legalised, as this would destigmatise it, suggesting, to some extent, they navigate the stigma they face as a result of their illicit cannabis use. However, other non-users accepted their friends’ cannabis use, so long as they were not directly exposed to it.

Most of our participants used cannabis as part of a specific group where they felt comfortable and confident in their control (Goffman, 1959). Cannabis effects were dependent on the quantity and the potency of cannabis. The effects ranged from feeling more relaxed physically, reducing anxiety and negative emotions, in line with a ‘chilled’ persona, and enabling adolescents to open-up to others (Lamb, 2011; Menghrajani et al., 2005). They reported that hash may be more conducive to feeling relaxed, whilst high doses of skunk could produce psychotic effects. Counterintuitively, preferences for skunk were expressed.

Both groups were aware of the potential risks of cannabis, unlike previous qualitative research which had suggested that adolescents minimised harmful consequences (Sandberg, 2012). However, adolescents seemed to enjoy talking about their experiences of negative effects and demonstrating their expertise about the type of cannabis they use.

Our research provided evidence that adolescent users rarely view their own cannabis use as addictive or problematic. Rather than seeking ‘deviant’ or addicted identities, participants described a range of social benefits, and specified that they did not need to use cannabis, they chose it. Their autonomy was also demonstrated in showing expertise in the more risky aspects of cannabis use. This reflects wider developmental processes during the adolescent identity transition, where parental control is lost and teenagers enjoy greater autonomy (Lamb, 2011). Therefore, while cannabis was not reported to be central in adolescents’ understandings of themselves, it may meet some of their social and developmental needs at a time of identity transition.

This study has some limitations. The focus groups were relatively small in size. The non-user group was not balanced in terms of gender, including one male and five females. Future research could aim for more representative and larger samples. Also, whilst data collection was anonymous, adolescents may be subjected to social desirability bias, because they completed it in front of others. Future work should expand research into other geographical areas and cultures.

## Conclusion

This study examined 16-17 year-old London based adolescents thoughts and attitudes about cannabis and identity. Despite acknowledging its harmful consequences, adolescents stated that cannabis makes them relaxed and provides pleasure. Cannabis users were seen as ‘chilled’ and they valued their expertise in risky activities. Users did not explicitly think that cannabis is part of their self-identity, despite evidence suggesting they publicise and defend their own cannabis use. To the adolescents who used cannabis, the drug plays a role in their lives as something that they *do*, more than something that they *are*. Some non-users held negative, devaluing stereotypes of people who use cannabis. Moralistic, prejudiced and stigmatising views are still held about adolescents who use cannabis, by some of their non-using peers. Efforts to de-stigmatise adolescent cannabis use could contribute to honest, drugs education; harm reduction advice; and increase acceptability of treatment for those who use problematically.

## Data Availability

All data produced in the present study are available upon reasonable request to the authors.

## Acknowledgements

We thank all the adolescents who participated in this research project.

